# Associations of reading skills and properties of cerebral white matter pathways in 8-year-old children born preterm

**DOI:** 10.1101/2020.12.11.20247965

**Authors:** Sarah E Dubner, Michal Ben-Shachar, Aviv Mezer, Heidi M Feldman, Katherine E Travis

**Author notes:** **Corresponding Author:** Sarah E. Dubner, MD, 1265 Welch Rd. x109, Stanford, CA 94305.

## Abstract

**AIM:** Children born preterm (PT) experience perinatal white matter injury and later reading deficits at school age. We used two complementary neuroimaging modalities to determine if reading skills would be associated with contemporaneous white matter properties in school-aged PT children.

**METHOD:** In 8-year-old PT children (N=29), we measured diffusivity (fractional anisotropy, FA), from diffusion MRI, and myelin content (relaxation rate, R1) from quantitative relaxometry. We assessed reading (Gray’s Oral Reading Test, Fifth Edition) in each child. Whole-brain deterministic tractography coupled with automatic segmentation and quantification were applied to extract FA and R1 along four tracts and assess their statistical association with reading scores.

**RESULTS:** Reading-FA correlations were not significant along the four analyzed tracts. Reading-R1 correlations were significantly positive in portions of the left superior longitudinal fasciculus, right uncinate fasciculus, and left inferior longitudinal fasciculus. FA positively correlated with R1 in limited areas of reading-R1 associations, but did not contribute to the variance in reading scores.

**INTERPRETATION:** Combining complementary neuroimaging approaches identified relations between reading and white matter properties not found using a single MRI measure. Associations of reading skills and white matter properties may vary across white matter tracts and metrics in PT children.

**What this paper adds:**

▪ Preterm children’s reading was associated with white matter myelin content.
▪ Preterm children’s reading was not associated with white matter diffusivity.

---

Children born very or extremely preterm (PT), at < 32 weeks’ gestation, are at risk for reading problems (1). Deficits have been attributed to cerebral white matter injury and dysmaturity, part of a complex of encephalopathic changes incurred at birth (2). White matter differences consistent with encephalopathy of prematurity can be identified on neonatal diffusion magnetic resonance imaging scans (dMRI) as decreased directional diffusivity (fractional anisotropy, FA) or increased mean diffusivity (3). Differences in dMRI white matter metrics persist from childhood (4) through adulthood (5). White matter changes in PT children are thought to impact neurocognitive functions, such as reading, by altering structural and functional connectivity (6). However, studies of the associations of reading and white matter metrics in PT children show inconsistent findings (7). Identifying a neurobiological explanation for variability in findings is challenging because FA is influenced by multiple tissue properties (8). Complementary imaging methods have been used to interpret dMRI metric associations with neurocognitive outcomes in children born full term (FT) (9). Here we apply this novel approach to understanding the neurobiological basis of reading skills in PT children.

FT children’s reading skills are associated with white matter microstructural metrics derived from dMRI (10,11). Associations in dorsal pathways, including the arcuate fasciculus (Arc) and superior longitudinal fasciculus (SLF) (7,10–12), are thought to relate to transmission of phonological information for converting visual word forms to spoken words (13). Associations in ventral pathways, including the inferior longitudinal fasciculus (ILF) and the uncinate fasciculus (UF) are thought to relate to transmission of semantic and orthographic information (7,12,14,15). Individual differences in reading among PT children have also been associated with white matter microstructural metrics (16–18). However, different patterns of association of reading and white matter microstructural metrics have been found in PT versus FT children (7). The findings suggest that a different balance of white matter properties may lead to individual differences in reading abilities in PT versus FT children.

Quantitative relaxometry (qR) is an alternative MRI technique that can be used to assess white matter microstructure. qR measures the longitudinal relaxation rate of water protons in a magnetic field (R1, seconds^-1^). R1 is highly correlated with myelin content in white matter areas (19). Developmental changes identified by R1 are presumed to reflect maturational increases in myelin and related molecular factors (20– 22). Structural differences measured by R1 in PT and FT children are presumed to reflect primarily differences in the amount of myelin (23). qR metrics have been related to reading in typically developing FT children (9,24). R1 has not been used to examine individual variations in neurocognitive abilities in PT children. Further, studies have yet to combine both dMRI and qR to assess the contributions of these metrics in explaining variation in reading skills in PT children.

The present study assessed a sample of PT children who were part of a longitudinal study of the neural bases of reading development (15). When these children turned age 8 years, qR became available at our imaging center. To interrogate tissue properties associated with reading in this PT sample, we collected both dMRI and qR in these children. The aims of this study were: (1) To assess the associations between reading skills and FA of reading-related dorsal and ventral white matter pathways. (2) To assess the associations between reading skills and R1 in the same pathways. We hypothesized that we would not observe associations between reading skills and FA (12). We hypothesized that we would observe positive associations between reading skills and R1, given group differences in R1 in a PT and FT sample (23) and the assumption that these the group differences impact function. We assessed the degree of associations between FA and R1 in regions of significant association of reading and one or both white matter metrics and the relative contributions of each measure to reading scores.

## METHOD

### Participants

8-year-old PT children (*n*=43) were recruited between 2012 to 2015 as part of a longitudinal study (15). The present study included participants with useable dMRI and qR data and complete cognitive and reading measures. 14 participants were excluded because they underwent a different MRI protocol (n=2), did not undergo qR imaging (n=9), or showed excessive motion (n=3). The final sample consisted of 29 participants. We did not include a FT comparison group because we have previously documented differences in reading-FA associations (12).

PT birth was defined as gestational age (GA) *≤* 32 weeks. Below this GA, neonates are at greatest risk for white matter injury (2). English-speaking children were recruited from the Lucile Packard Children’s Hospital Stanford High-Risk Infant Follow-Up Clinic, local parent groups, and the surrounding San Francisco Bay Area. Exclusion criteria included neurological factors unrelated to prematurity that would account for white matter differences or impact learning to read English, including congenital anomalies, active seizure disorder, hydrocephalus, sensorineural hearing loss, visual impairment, intelligence quotient <80. The Stanford University Institutional Review Board approved the experimental protocol (#IRB-22233). A parent or legal guardian provided informed written consent. Children provided assent at age 8. Participants were compensated for participation.

Demographic characteristics included socio-economic status, measured using a modified 4-Factor Hollingshead Index (25). Handedness information was measured by the Edinburgh Handedness Inventory (26). Detailed medical information was available for 26 of 29 participants. A neuroradiologist assessed T1w scans collected as part of the current study for 5 features associated with white matter injury (15).

### Neurocognitive Measures

Reading ability measures were obtained as part of a battery of norm-referenced behavioral tests (12,15,18). We analyzed the Oral Reading Index (ORI) generated from the Gray’s Oral Reading Tests, 5^th^ Edition (27). The ORI, a composite score composed of fluency (rate and accuracy) and comprehension, is reported as a standard score (mean=100, standard deviation=15). General intelligence was assessed using the Wechsler Abbreviated Scale of Intelligence (WASI-II) (28). Study assessments were completed in English.

### MRI Acquisition and Analysis

MRI data were acquired on a 3T Discovery MR750 scanner (General Electric Healthcare, Milwaukee, WI, USA) equipped with a 32-channel head coil (Nova Medical, Wilmington, MA, USA) at the Center for Cognitive and Neurobiological Imaging at Stanford University (www.cni.stanford.edu). Participants were scanned without sedation. High-resolution T1-weighted (T1w) anatomical images were collected using an inversion recovery (IR)-prep 3D fast-spoiled gradient-echo (FSPGR) sequence collected in the sagittal plane (0.9 x 0.9 x 0. 9 mm^3^ voxel size). The T1w image was used as an anatomical reference for the alignment of the diffusion tensor image (DTI) maps.

dMRI data were acquired with a diffusion-weighted, dual-spin echo, echo-planar imaging sequence with full brain coverage. Diffusion weighted gradients were applied at 30 non-collinear directions with a b-value=1,000 s/mm^2^. Three volumes were acquired at *b*=0 at the beginning of the scan. We collected 70 axial slices in each participant (TR=8300 ms; TE=83.1 ms; FOV=220 mm; matrix size of 256 x 256, 0.8594 x 0.8594 x 2 mm^3^ voxel size).

qR relaxometry was acquired with SPGR echo images acquired at four flip angles (*α* = 4°, 10°, 20°, and 30°; TR=14 ms; TE=2 ms) (29). SPGR scans were acquired sequentially, without breaks to ensure that scan parameters were similar across flip angles. A 0.9375 x 0.9375 x 1.5 mm^3^ voxel size was chosen to acquire the four SPGR scans in a reasonably short time frame, appropriate for a pediatric clinical sample. To obtain an unbiased R1 map, we acquired a spin-echo inversion-recovery (SEIR) sequence with echo-planar imaging (EPI) read-out (30) measured at lower resolution (1.875 x 1.875 x 4 mm^3^ voxel size). Four inversion times were measured (2400, 1200, 400 and 50 ms; TR = 3 s; TE = 47 ms). Unbiased R1 maps obtained from SEIR-EPI scans were used to correct for RF transmit bias in the higher-resolution SPGR scans as described and validated by (31).

Image preprocessing, registration, deterministic tractography, fiber tract identification, segmentation and quantification are described in the **Supplemental Information**. We computed fractional anisotropy (FA) from dMRI scans and derived R1 from qR scans. The validity of the current methods have been demonstrated in developmental studies (20). Additionally, evidence shows reduced R1 in patients with clinical conditions in which myelin content is decreased (23,31). We characterized white matter metrics from two dorsal tracts (Arc-L and bilateral SLF) and two bilateral ventral white matter tracts (ILF and UF). We selected these pathways *a priori* based on evidence implicating their involvement in phonological and semantic processes (13,32).

**Figure 1** shows the left hemisphere tracts in a representative participant. We generated tract profiles that measured FA or R1 at 30 equidistant locations along the central portion of each fiber tract, bounded by the same ROIs used for tract segmentation. We averaged FA or R1 values across all 30 locations to generate a mean metric for each tract.

**Figure 1.**
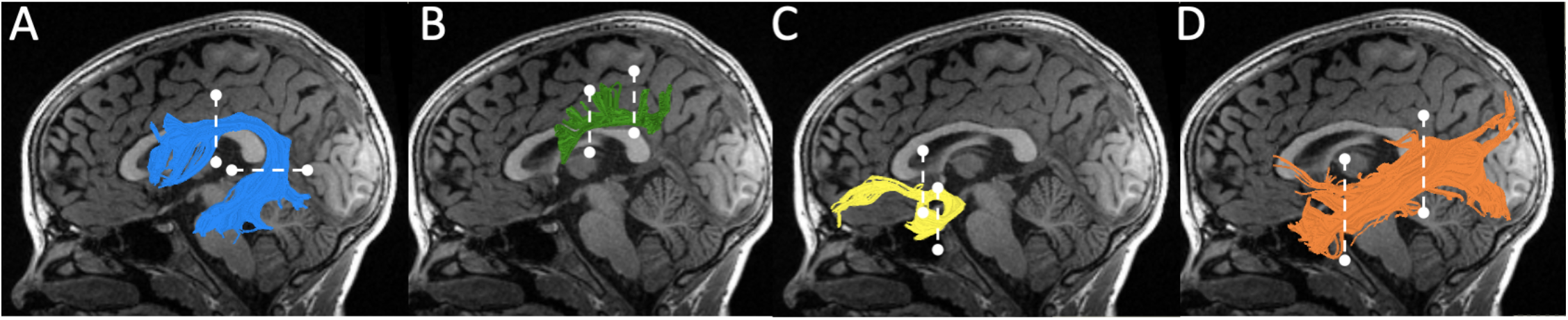
Tractography of selected white matter tracts. Left hemisphere tract renderings are displayed on a mid-sagittal T1 image from a representative participant. Right hemisphere tract renderings not shown. A) The arcuate fasciculus is shown in blue, B) the superior longitudinal fasciculus is shown in green, C) the uncinate fasciculus is shown in yellow, D) the inferior longitudinal fasciculus is shown in orange. Dashed lines represent the location of the regions of interest (ROIs) used to segment each pathway from the whole-brain tractogram.

### Statistical Approach

Statistical analyses were conducted using SPSS (version 26.0, IBM Corp., 2019). Statistical significance was set at *p*<0.05. Normality of clinical and neurobiological data was assessed using the Shapiro–Wilk test. We used parametric tests for all associations. A False Discovery Rate (FDR) of 5% was calculated for zero-order associations to account for multiple comparisons (33).

### Associations between reading and white matter microstructure metrics

We computed Pearson correlations to assess associations between ORI and mean FA or mean R1. To identify relations obscured by mean measures, we computed correlations between ORI and FA or R1 for 30 equidistant nodes along each tract. The tract profile analysis utilized a nonparametric permutation-based method to control for 30 along-tract comparisons (34), producing a family-wise error cluster size and a critical r-value for each tract. Along-tract correlations were considered significant if (1) correlations at an uncorrected level of p<0.05 occurred in adjacent nodes *≥* the critical cluster size (range 6-9 nodes) or (2) correlations *≥* the critical r-value occurred within a cluster size *≥* 3 nodes. For FA and R1, we computed the mean r-value from the longest tract segment meeting these criteria and applied a false discovery rate (FDR) at p<0.05, to control for multiple comparisons across 7 tracts.

In order to understand whether myelin was the dominant tissue property represented by FA in regions of reading-white matter metric correlations, we computed Pearson correlations of mean FA and mean R1 for the segments with significant reading-white matter correlations. We also conducted a series of multiple linear regression models to assess the unique contributions of mean R1 and mean FA to ORI scores in the segments along each tract with significant reading-white matter correlations.

## RESULTS

### Participant Characteristics

Participant characteristics are shown in **Table 1**. Four of 29 participants had mildly abnormal T1w scans. Mean reading and IQ scores fell within the normal range.

**Table 1.** Participant Characteristics.

|  | **N = 29** |
| --- | --- |
| **Demographic measures** | **Mean (sd)** |
| Age at Testing, Years | 8.2 (0.2) |
| Birthweight, Grams | 1279 (419) |
| Gestational Age, Weeks | 29.2 (2.2) |
| Male, n (%) | 17 (58.6) |
| SES<sup>a</sup> | 56.9 (9.7) |
| Left-Handed, n (%) | 2 (7.9) |
| **Neonatal Medical Complications (n = 26)** |  |
| Small for Gestational Age, n | 2 |
| Respiratory Distress Syndrome, n | 21 |
| Bronchopulmonary Dysplasia, n | 4 |
| Hyperbilirubinemia, n | 20 |
| Patent Ductus Arteriosus, n | 11 |
| Retinopathy of Prematurity or Immature Retinae, n | 15 |
| Necrotizing Enterocolitis, n | 4 |
| **Neonatal Head Ultrasound or MRI findings (n = 26)** |  |
| Grade I Intraventricular hemorrhage (IVH) | 5 |
| Grade II IVH | 1 |
| Grade III IVH | 1 |
| Small periventricular lesions | 1 |
| Mild white matter injury | 3 |
| Transient vascular malformation | 1 |
| Enlarged ventricles | 3 |
| **Neurocognitive Measures** |  |
| Oral Reading Index<sup>b</sup> | 95.9 (12.0) |
| Full 4 IQ Scaled Score<sup>c</sup> | 105.5 (12.5) |

<sup>a</sup> SES, Socioeconomic Status, Modified Hollingshead Index.
<sup>b</sup> Oral Reading Index Scaled Score, Gray Oral Reading Tests, 5<sup>th</sup> Edition.
<sup>c</sup> Wechsler Abbreviated Scale of Intelligence, 2<sup>nd</sup> Edition, Full 4.

### Reading and FA

Mean FA values are shown in **Supplemental Table S1**. No significant correlations were identified between reading and mean FA in any tract (**Table 2)**. Reading and along-tract FA correlations confirmed that mean measures did not obscure significant associations in any tract.

**Table 2.** Correlations among mean-tract FA, mean-tract R1, and Reading.

| Tract | Pearson Correlation (r) |  |  |  |
| --- | --- | --- | --- | --- |
|  | FA*Reading |  | R1*Reading |  |
| **Dorsal** | **r** | **p** | **r** | **p** |
| Arc-L | -0.003 | 0.99 | 0.21 | 0.27 |
| SLF-L | 0.03 | 0.89 | 0.36 | 0.06 |
| SLF-R | -0.28 | 0.14 | -0.15 | 0.43 |
| **Ventral** |  |  |  |  |
| UF-L | 0.18 | 0.35 | 0.32 | 0.09 |
| UF-R | 0.25 | 0.18 | 0.34 | 0.11 |
| ILF-L | -0.18 | 0.34 | 0.35 | 0.06 |
| ILF-R | -0.07 | 0.72 | 0.12 | 0.54 |

Arc, Arcuate fasciculus; SLF, Superior longitudinal fasciculus; ILF, Inferior longitudinal fasciculus; UF, Uncinate fasciculus; Reading, Oral Reading Index.

### Reading and R1

Mean R1 values are shown in **Supplemental Table S1**. Moderate positive correlations were found between reading and mean R1 in the SLF-L, UF-L, UF-R, and ILF-L (**Table 2)**, but those did not reach statistical significance. Along-tract analysis revealed significant positive correlations between reading and R1 in the SLF-L (nodes 1-5; r_mean_=0.42), UF-R (nodes 13-19; r_mean_=0.45), and ILF-L (nodes 20-23 r_mean_=0.53) (p<0.05, FWE corrected for 30 nodes within each tract), as shown in **Figure 2**. These associations remained significant (p<0.05) after controlling for the FDR across 7 tracts. *FA and R1*

**Figure 2.**
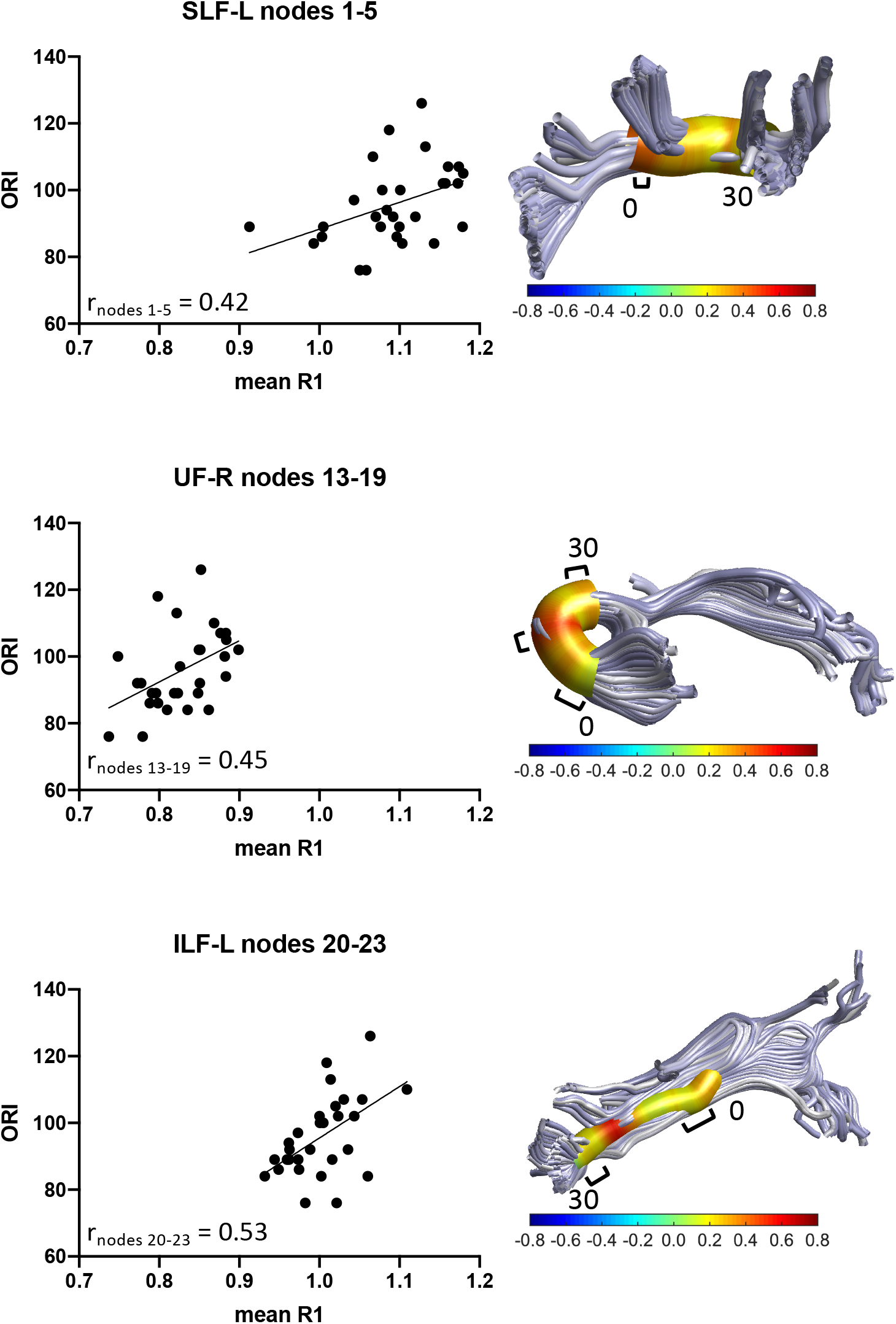
Associations between along-tract R1 and ORI at age 8 years in PT children. Each panel includes a rendering of a specific tract with the strength of the Pearson correlation (r) between R1 and ORI at 30 equidistant nodes along the tract represented as a heat-map cylinder surrounding the tract. Each panel also includes a scatter plot representing the strength of the association between mean R1 of the adjacent nodes in the longest region of the tract with significant correlations between R1 and ORI. Panels show the results for the SLF-L, UF-R, and ILF-L. R1 is significantly positively associated with reading within regions of these tracts. Brackets ([) indicate approximate regions of the tract with significant positive correlations between FA and R1. R1 = Relaxation rate (s^-1^); ORI = Oral Reading Index; SLF-L = Left Superior Longitudinal Fasciculus; UF-R = Right Uncinate Fasciculus; ILF-L = Inferior Longitudinal Fasciculus.

We calculated the strength of association between mean FA and mean R1 of the nodes in the segments of the SLF-L, UF-R, and ILF-L where reading was significantly associated with R1 (**Supplemental Figure S1)**. In the SLF-L, the FA-R1 associations were significant in the region of R1-reading association (r = 0.52). Correlations between mean FA and mean R1 in the UF-R and ILF-L did not reach statistical significance. In separate multiple linear regression models we confirmed that R1 made a significant contribution to ORI all three tracts - SLF-L R^2^ 15% (p = 0.022), UF-R R^2^ 17% (p = 0.017), ILF-L R^2^ 25% (p = 0.003) (**Supplementary Table 2**). FA did not contribute unique variance in any of the models.

## DISCUSSION

In this investigation of 8-year-old PT children, we interrogated the associations of reading skills and concurrent white matter microstructure metrics from two complementary imaging methods, dMRI and qR. We did not find any associations between reading and FA, consistent with previous findings. A novel finding was that we found significant associations between PT children’s reading abilities and R1, a measure from qR imaging that is highly sensitive to myelin content within white matter tracts. The location of the positive associations included a dorsal tract (SLF-L) and two ventral tracts (UF-R, and ILF-L) that have been implicated in reading in many studies of FT children. We found that significant positive associations between FA and R1 localized to the region of reading-R1 associations within the SLF-L, but not within the UF-R or ILF-L. However, in the SLF-L, FA did not make a unique contribution to the variance in reading scores. These findings contribute substantially to understanding the reading outcomes of preterm birth at school age.

### Correlations between reading and white matter metrics

We did not observe associations between reading skills and FA within any of the tracts. This finding is consistent with our previous work that found FA at age 6 was related to reading skills at both age 6 and 8 years in FT children but not in PT children (12,15). The findings suggested that individual differences in reading among PT children are not associated with directional diffusivity at this age. However, positive associations between reading skills and FA have been found at older ages in PT children (7).

In this sample of PT children, R1 proved to be more sensitive to individual variation in reading than did FA. R1 within white matter tracts is considered a proxy for myelin content of the tract. Long-term neurodevelopmental outcomes in PT children are currently thought to result from injury and/or aberrant regeneration and repair of oligodendrocyte precursor cell injury that are part of the encephalopathic changes of PT birth. R1 may be more directly associated with these early processes than is FA. Generally, myelin is thought to contribute to directional diffusion by restricting water diffusion in directions perpendicular to the main direction of the axon. However, other factors also contribute to FA, including axonal diameter, fiber coherence, and proportion of crossing fibers. It is possible that these other factors have a greater impact on FA or neutralize the influence of myelin content on FA in PT children, thereby reducing the strength of reading-FA associations. Alternatively, part of the R1 effect could be related to iron or lipid content that effect R1 and not FA (21,22).

### Tracts in which associations were found

The location of reading-R1 associations is consistent with studies of white matter microstructural metrics derived from diffusion studies in FT children. Several reading skills have been associated with SLF white matter microstructural metrics in samples of FT children (7,12). Reading comprehension has been associated with UF white matter microstructural metrics (14). In 6-year-old FT children, reading measures have been associated with FA of both the UF-R and SLF-L, but associations were not found in these tracts in the present PT cohort at age 6 (15,18). ILF maturation relates to children’s reading abilities (12) and ILF-L microstructural metrics have been associated with reading comprehension in FT children with weaker decoding skills (14). Here, associations between reading and R1 of these tracts in PT children, suggest that these tracts are also important for reading in PT children. However, the white matter microstructural characteristics related to reading in PT children may be different from those in FT children.

### FA-R1 associations

Previous studies of FT children or adults have shown FA to be predominately affected by fiber crossings (35) with minimal correlations between FA and qR metrics (36). In PT children the sequelae of prematurity may disrupt the normal progression of myelination and axonal development, potentially increasing variability in the tissue composition of myelin and other axonal properties in PT children (2). Here, FA and R1 were not consistently correlated in areas of reading-R1 correlation, highlighting the complexity of reading-white matter associations. Likely, not all white matter tracts operate in the same manner in the service of reading. Additionally, white matter properties may not be equally represented by FA and R1 across tracts. Other properties, such as axon diameter, may contribute to FA. Longitudinal studies are needed to clarify how white matter maturation and prematurity affect the balance of tissue properties underlying variations in dMRI metrics throughout childhood. After accounting for the contribution of myelin to variance in reading, FA did not explain significant additional variance in reading in this sample. Future studies combining analytic approaches for measuring fiber dispersion and packing from dMRI (37) with structural MRI techniques for assessing axonal diameter (38) or g-ratio (22) will aid in understanding how myelin and other axonal properties relate to variability in FA and reading within and across clinical populations.

Limitations include a modest sample size and reading scores within the normal range. Future studies with larger samples with greater diversity in reading skills could confirm the findings described here and explore whether associations exist in other white matter pathways. dMRI with increased number of directions would allow use of probabilistic tractography.

## Conclusions

In summary, by combining complementary structural neuroimaging approaches we identified relations between reading and concurrent white matter properties that were not found using a single measure from dMRI in isolation. Variation in relations between FA and R1 in regions of significant reading-R1 correlation suggested that the white matter properties represented by FA vary by fiber group and that myelin plays an important role in reading abilities in PT children compared to other white matter properties. Further exploration of white matter pathways using multiple imaging techniques will contribute to understanding reading-brain associations in the aftermath of preterm birth.

## Data Availability

Anonymized data will be made available upon request to qualified investigators for purposes of replicating procedures and results from the corresponding author.

## ABBREVIATIONS

AFQ: Automated Fiber Quantification
Arc-L: left arcuate fasciculus
Arc-R: right arcuate fasciculus
dMRI: diffusion magnetic resonance imaging
DTI: Diffusion Tensor Imaging
FSPGR: fast-spoiled gradient-echo
FA: fractional anisotropy
FT: full term
GA: gestational age
ILF-L: left inferior longitudinal fasciculus
ILF-R: right inferior longitudinal fasciculus
MRI: magnetic resonance imaging
ORI: Oral Reading Index
PT: preterm
qR: Quantitative Relaxometry
R1: relaxation rate (seconds^-1^)
ROI: region of Interest
SES: socioeconomic status
SLF-L: left superior longitudinal fasciculus
SLF-R: right superior longitudinal fasciculus
T1w: T1-weighted
UF-L: left uncinate fasciculus
UF-R: Right uncinate fasciculus
WASI-II: Wechsler Abbreviated Scale of Intelligence, 2^nd^ Edition.

## Acknowledgments

The authors received funding support from the National Institute of Child Health and Human Development (Grant # R01HD069162 [Feldman], 5K99HD084749 [Travis]), a Young Investigator Award to Dr. Travis, from the Society of Developmental and Behavioral Pediatrics (2014), and a Tashia and John Morgridge endowed fellowship through the Stanford Maternal Child Health Research Institute Clinical Trainee Grant to Dr. Dubner. Funders were not involved in study design, data collection, data analysis, manuscript preparation or publication decisions.

