## Supplemental Information for "Associations of reading skills and properties of cerebral white matter pathways in 8-year-old children born preterm"

**Image Preprocessing:** The T1w images were first aligned to the canonical ac-pc orientation. Diffusion weighted images were pre-processed with Vistasoft (http://github.com/vistalab/vistasoft/mrDiffusion), an open-source software package implemented in MATLAB R2012a (Mathworks, Natick, MA). The dual-spin echo dMRI sequence used here greatly reduces eddy-current distortions obviating the need for eddy-current correction (1). Subject motion during the diffusion weighted scan was corrected using a rigid body alignment algorithm (2). Each diffusion weighted image was registered to the mean of the $b$=0 images and the mean $b$=0 image was registered automatically to the participant's T1w image, using a rigid body transformation (implemented in SPM8, http://www.fil.ion.ucl.ac.uk/spm/; no warping was applied). The combined transform that resulted from motion correction and alignment to the T1w anatomy was applied to the raw data once, and the transformed images were resampled to 2 x 2 x 2mm$^3$ isotropic voxels. This step was performed because non-isotropic voxels may bias the tensor fit and distort both tracking and measurements of diffusion properties (3). Diffusion gradient directions were then adjusted to fit the resampled diffusion data (4).

For each voxel in the aligned and resampled volume, tensors were fit to the diffusion measurements using a robust least-squares algorithm, Robust Estimation of Tensors by Outlier Rejection (RESTORE), which is designed to remove outliers at the tensor estimation step (5). A continuous tensor field was estimated using trilinear interpolation of the tensor elements. The eigenvalue decomposition of the diffusion tensor was calculated and the resulting three eigenvalues ($\lambda$1, $\lambda$2, $\lambda$3) were used to compute fractional anisotropy (FA), mean diffusivity (MD; i.e., the mean of $\lambda$1, $\lambda$2,and

$\lambda 3$), radial diffusivity (RD, i.e., the mean of $\lambda 2$ and $\lambda 3$) and axial diffusivity (AD, i.e., $\lambda 1$) (6).

Quantitative T1 (relaxation time, seconds) maps were calculated using mrQ, (https://github.com/mezera/mrQ), an open-source software package implemented in MATLAB R2012a (Mathworks, Natick, MA). T1 fitting and bias correction was calculated using methods described in (7,8). These procedures involve measuring the local transmit-coil inhomogeneities which are calculated by minimizing the difference between the unbiased T1 map calculated from the low-resolution SEIR-EPI images (9) and the T1 map fit from the high-resolution multiple flip angle SPGR images. Minimization was achieved using a nonlinear least-squares (NLS) solution that assumes transmit-coil inhomogeneities to be smooth in space (10).

To co-register a subject’s quantitative relaxometry (qR) images to their dMRI data, we used the Advanced Normalization Tools (ANTs) software package (11). This tool was used to warp the qR images to the non-diffusion weighted b=0 images, as these images have relatively similar contrast (12). This warping procedure is used to minimize mis-registration errors due to EPI distortions in the dMRI data. EPI distortions were minimal due to the 2x ASSET acceleration used for the readout of the diffusion-weighted images. After applying the diffeomorphic warp, image registration was manually inspected using the mrView software (13). Manual inspection of the aligned images confirmed that the registration was accurate in all subjects. From the qR maps, we then derived R1, the inverse of T1 (R1 = 1/T1).

***Quantification of White Matter Tissue Properties***

Automated Fiber Quantification (AFQ; https://github.jyeatman/AFQ; (14)), a software package implemented in MATLAB R2012a (Mathworks, Natick, MA), was used to isolate and characterize white matter metrics from two dorsal tracts: the Arc-L and bilateral SLF, and two bilateral ventral white matter tracts: the ILF and UF. We selected these pathways *a priori* based on evidence implicating their involvement in phonological and semantic processes (15–18). Due to previously described limitations of deterministic tractography for segmenting the right Arc (19–23), we excluded it from analysis *a priori*. Using AFQ software, we generated an FA or R1 tract profile that described the variations in either tissue parameter (FA and R1) along the central portion of the tract. Specifically, tract FA or tract R1 profiles each were calculated at 30 equidistant locations along the central portion of each fiber tract bounded by the same two ROIs used for tract segmentation. The core of the tract was calculated by defining 30 sample-points along the tract and computing the robust mean position of the corresponding sample points. The robust mean for all tracts except the SLF was computed by estimating the 3-dimensional Gaussian covariance of the sample points and removing fibers that are either located more than 5 standard deviations away from the mean position of the tract, or that differed more than 4 standard deviations in length from the mean length of the tract. In the SLF, these parameters resulted in the inclusion of many fibers not consistent with the known anatomy of the SLF. Therefore, for the SLF, fibers that were 4 standard deviations away from the mean position of the tract, or that differed more than 1 standard deviation in length from the mean length of the tract were removed. This computation constituted the final automatic cleaning stage of the segmented tracts. Tract profiles were then averaged to produce a single mean FA or R1 value for each tract. Importantly, the FA and R1 measurements were obtained for the same tract regions.
