## Supplemental Figure S1 for "Associations of reading skills and properties of cerebral white matter pathways in 8-year-old children born preterm"

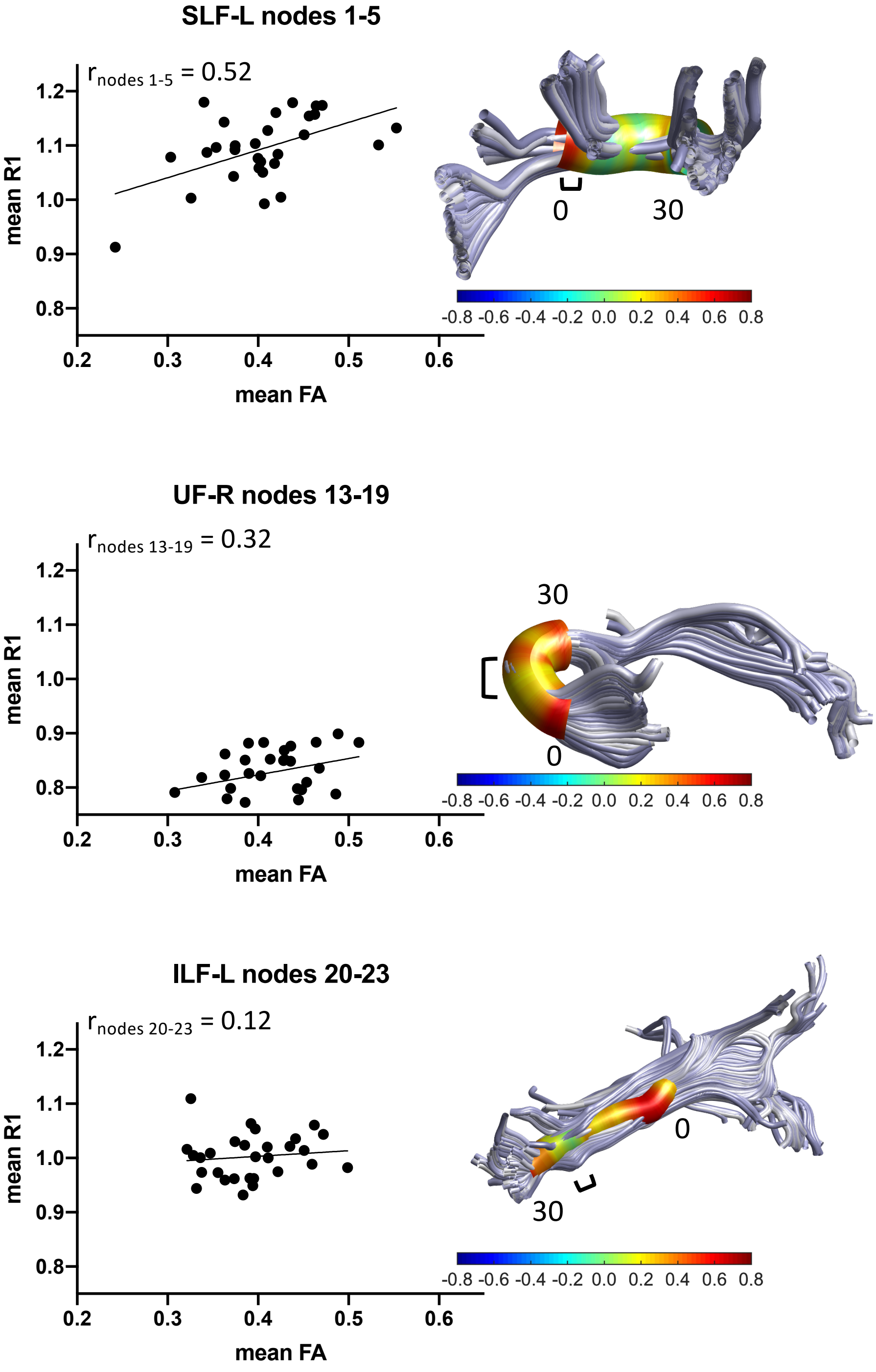


**Figure S1. Associations between FA and R1 at age 8 years in children born preterm**. Each panel includes a rendering of a specific tract with the strength of the Pearson correlation (r) between FA and R1 at 30 equidistant nodes along the tract represented as a heat-map cylinder surrounding the tract. Each panel also includes a scatter plot representing the strength of the association between mean FA and mean R1 of the contiguous nodes in the region of the tract with significant correlations between ORI and R1. Brackets ( **[** ) indicate approximate regions of the tract with significant positive correlations between ORI and R1.
FA = fractional anisotropy; R1 = Relaxation rate ($s^{-1}$); SLF-L = Left Superior Longitudinal Fasciculus; UF-R = Right Uncinate Fasciculus; ILF-L = Left Inferior Longitudinal Fasciculus.
