## Supplemental Table S1 for "Associations of reading skills and properties of cerebral white matter pathways in 8-year-old children born preterm"

**Supplemental Table S1. Mean FA and R1**

| Tract | Mean FA | Mean R1 |
|---|---|---|
| | Mean (SD) | Mean (SD) |
| **Arc-L** | 0.48 (0.03) | 1.12 (0.03) |
| **SLF-L** | 0.43 (0.04) | 1.11 (0.05) |
| **SLF-R** | 0.49 (0.05) | 1.10 (0.05) |
| **UF-L** | 0.42 (0.03) | 0.83 (0.05) |
| **UF-R** | 0.43 (0.04) | 0.85 (0.05) |
| **ILF-L** | 0.45 (0.04) | 1.03 (0.04) |
| **ILF-R** | 0.45 (0.04) | 1.12 (0.03) |

Arc, Arcuate fasciculus; SLF, Superior longitudinal fasciculus; ILF, Inferior longitudinal fasciculus; UF, Uncinate fasciculus.
