## Supplemental Table S1 for "Associations of reading skills and properties of cerebral white matter pathways in 8-year-old children born preterm"

**Table S2.** Multiple linear regression models assessing the unique contribution of R1 and FA to reading outcome (Oral Reading Index) at age 8 years in A. the left superior longitudinal fasciculus (SLF-L). B. the right uncinate fasciculus (UF-R), and C. the left inferior longitudinal fasciculus (ILF-L).

| **A. SLF nodes 1-5** | **Model A1** | **Model A2** |
|---|---|---|
| **R1** | 0.42 | 0.39 |
| **FA** | | 0.068 |
| **$\Delta$ $R^2$** | | 0.003 (p = 0.74) |
| **Total $R^2$** | 0.18 (p = 0.02) | 0.18 (p = 0.07) |
| **Adj $R^2$** | 0.15 | 0.12 |
| **B. UF-R nodes 13-19** | **Model B1** | **Model B2** |
| **R1** | 0.45 | 0.40 |
| **FA** | | 0.17 |
| **$\Delta$ $R^2$** | | 0.02 (p = 0.37) |
| **Total $R^2$** | 0.20 (p = 0.015) | 0.23 (p = 0.04) |
| **Adj $R^2$** | 0.17 | 0. 17 |
| **C. ILF-L nodes 20-23** | **Model C1** | **Model C2** |
| **R1** | 0.53 | 0.56 |
| **FA** | | -0.31 |
| **$\Delta$ $R^2$** | | 0.09 (p = 0.06) |
| **Total $R^2$** | 0.28 (p = 0.003) | 0.37 (p = 0.002) |
| **Adj $R^2$** | 0.25 | 0. 32 |

Data are standardized coefficients. $R^2$ change (**Δ** $R^2$) values in Model 2 reflect the increase in variance accounted for by FA in addition to R1 (Model 1).
